# Cost-Effectiveness Analysis of Statins, Berberine, and Their Combined Use for Primary Prevention of Cardiovascular Disease

**DOI:** 10.1101/2025.02.20.25322455

**Authors:** Yuanqing Xia, Kathy Leung, Jie V. Zhao

**Affiliations:** School of Public Health, Li Ka Shing Faculty of Medicine, The University of Hong Kong, Patrick Manson Building, 7 Sassoon Road, Hong Kong SAR, P. R. China; The Hong Kong Jockey Club Global Health Institute, Hong Kong Special Administrative Region, P. R. China; WHO Collaborating Centre for Infectious Disease Epidemiology and Control, School of Public Health, LKS Faculty of Medicine, The University of Hong Kong, Hong Kong SAR, P. R. China; Laboratory of Data Discovery for Health Limited (D24H), Hong Kong Science Park, Hong Kong SAR, P. R. China; The University of Hong Kong - Shenzhen Hospital, Shenzhen, P. R. China; State Key Laboratory of Pharmaceutical Biotechnology, The University of Hong Kong, Hong Kong SAR, China

**Keywords:** **Keyword**: cardiovascular diseases, cost-effectiveness, stains, berberine, primary prevention

## Abstract

**Background:** Statins are the cornerstone treatment for the primary prevention of cardiovascular disease (CVD), but intolerance and side effects can hinder adherence. Berberine, with promising lipid-lowering effects and good tolerance, presents a potential alternative for statin-intolerant patients.

**Aim:** To estimate and compare the cost-effectiveness of statins, berberine, and their combined use for primary CVD prevention.

**Methods:** The Scottish CVD Policy Model was used to predict long-term health and cost outcomes in Scottish adults aged 40 years or older without pre-existing CVD. Intervention and cost inputs were sourced from published literature and health service cost data. The primary outcome measure was the lifetime incremental cost-effectiveness ratio (ICER), evaluated as cost per quality-adjusted life year (QALY) gained. The intervention strategies of no intervention, atorvastatin 20 mg per day (“statins”), berberine 1000 mg per day (“berberine”), simvastatin 20 mg plus berberine 1500 mg per day (“combined intervention 1”) and simvastatin 20 mg plus berberine 900 mg per day (“combined intervention 2”) were analyzed for individuals with ASSIGN risk scores ≥20% and ≥10%.

**Results:** All intervention strategies were cost-effective (statins: ICER £1,260.7/QALY, 95%CI: £- 2,528.6/QALY ∼ £2,305.5/QALY; berberine: ICER £6,192.4/QALY, 95%CI: £4,655.7/QALY ∼ £11,387.0/QALY; combined intervention 1: ICER £5,506.5/QALY, 95%CI: £4,506.8/QALY ∼ £10,732.0/QALY; combined intervention 2: ICER £3,846.4/QALY, 95%CI: £3,107.0/QALY ∼ £5,270.1/QALY), compared to no intervention, at the threshold of ICER of £20,000 per QALY. Compared to statins, berberine was less cost-effective, but the combined interventions remained cost-effective (£10,198.6/QALY [95%CI: £6,740.4/QALY ∼ £58,473.3/QALY]; £6,362.8/QALY [95%CI: £5,187.7/QALY ∼ £12,499.2/QALY]) at the threshold of £20,000/QALY. Notably, when using drug costs from China (reflecting lower berberine prices), berberine and the combined interventions were preferable to statins alone.

**Conclusions:** Statins, berberine, and combined interventions are all cost-effective options for primary CVD prevention. Berberine could be considered a valuable alternative or complementary therapy, particularly if its price decreases below that of statins.

## 1. Introduction

Cardiovascular disease (CVD) is the leading cause of morbidity and mortality worldwide, which poses a heavy burden on healthcare.[1] The primary prevention of CVD through the management of dyslipidemia, a major risk factor for CVD, is a critical strategy for improving population health outcomes and reducing the associated economic impact. Statins (HMG-CoA reductase inhibitors) are lipid-lowering drugs used as a cornerstone treatment for the primary and secondary prevention of CVD.[2–4] Statin use has increased substantially in recent years,[5, 6] with nearly two-thirds of those taking statins for primary prevention.[7] The clinical guidelines also recommend statin uses among people previously considered at low risk of CVD.[4, 8–10] According to seven European Society of Cardiology/European Atherosclerosis Society (ESC/EAS) guidelines, the proportion of patients eligible for statins increased from approximately 8% in 1987 to 61% in 2016 due to the change in guidelines.[11] However, statin intolerance occurs in as many as 9.1% of patients treated with statins,[12] due to side effects, such as myopathy, hepatotoxicity and incident diabetes.[13–16]

In recent years, there has been growing interest in exploring natural, plant-derived compounds as alternative or complementary options for lipid-lowering.[17, 18] Berberine is one of the promising candidates. Several recent meta-analyses of randomized clinical trials (RCT) have provided compelling evidence for the lipid and lipoprotein-modulating effects of berberine alone and combined with statins.[19–21] Compared with the statins alone, berberine plus statins was more effective in lowering triglyceride and total cholesterol.[21] Previous studies also show that berberine is well-tolerated. Notably, unlike statins, which increase the risk of diabetes, berberine improves glucose metabolism.[22, 23] Given its favorable safety profile and lipid-lowering benefits, berberine has been proposed as a potential alternative therapy for treating dyslipidemia among patients with statin intolerance by both the International Lipid Expert Panel[24] and the 2019 ESC/EAS Guidelines[25].

However, the cost-effectiveness of berberine and berberine combined with statins is still unclear. We also did not identify any studies comparing berberine, berberine combined with statins versus statins in terms of overall cost-effectiveness. Considering the research gap, our study objective was to estimate and compare the cost-effectiveness of statins, berberine, and combined intervention of statins and berberine for primary prevention of CVD based on 10-year CVD risk.

## 2. Method

### 2.1 Study design

In this study, we estimated and compared the cost-effectiveness of different intervention strategies in a CVD-free population in the United Kingdom (UK) from a UK healthcare sector perspective. The intervention strategies include no intervention, statins, berberine, and the combination of statins with berberine for primary prevention of CVD. Individual eligibility for statins, berberine or combined interventions is determined based on each individual’s 10-year CVD risk, which was estimated using the ASSIGN score.[26]

#### 2.1.1 ASSIGN score

The ASSIGN score is primarily used in the UK, and it is recommended as the preferred risk assessment tool by the Scottish Intercollegiate Guidelines Network (SIGN) for cardiovascular risk assessment in the UK. It is designed to provide a more accurate assessment of CVD risk compared to other risk assessment tools, such as the Framingham risk score, by taking into account socioeconomic status as an independent risk factor.[27]

For individuals without pre-existing CVD, preventive statin therapy is prioritized based on their estimated 10-year risk of experiencing a primary CVD event, such as a heart attack or stroke.[28] The ASSIGN score is a cardiovascular risk assessment tool that is used to estimate an individual’s 10-year risk of developing CVD, expressed as a percentage.[26] It was developed by the SIGN using data from the Scottish Heart Health Extended Cohort (SHHEC). The ASSIGN score takes into account several risk factors, including age, sex, family history of premature CVD, smoking status, blood pressure, diabetes, total cholesterol (TC), high-density lipoprotein cholesterol (HDL-c) and Scottish Index of Multiple Deprivation (SIMD).

Initially, the 10-year CVD risk threshold for statin eligibility was set at 20%. However, in 2014, the National Institute for Health and Care Excellence (NICE) in England and Wales recommended lowering the risk threshold for statin initiation to a uniform 10%.[29] In contrast, the SIGN continues to uphold a threshold of 20%.[27] Given the varying thresholds set by different guidelines,[27, 29] our study used 10-year CVD risk thresholds of both 10% and 20% for intervention eligibility.

#### 2.1.2 Data Sources and intervention-eligible Population

We used data from the Scottish Health Survey (SHS) to simulate the CVD-free population, which are publicly available from the UK Data Service and can be accessed at https://beta.ukdataservice.ac.uk/datacatalogue/series/series?id=2000047. The SHS is a key national survey that collects comprehensive data on the health and wellbeing of the Scottish population. The SHS series was established in 1995, conducted annually with different focuses. The SHS collected data by questionnaire and nurse visit. The questionnaire covered information on individual’s demographic characteristics, lifestyle behaviors, and the prevalence of various health conditions. The nurse visits covered usage of medicines and supplements, physical examinations, blood sample, saliva sample and urine sample. Since the ASSIGN score implicated several risk factors, we finally included the SHS of 2003,2008,2009,2010 and 2011, which have collected blood samples or recorded systolic blood pressure.

We obtained eligible population for individuals aged 40 years or older without pre-existing CVD from SHS dataset. This aligns with the recommendations from the NICE in England and Wales, which advises initiating CVD risk assessment in the population 40 years and above.[28] Those currently receiving statins were also excluded. Demographic characteristics and CVD risk factor data were obtained from the SHS.

#### 2.1.3 The Scottish Cardiovascular Disease Policy Model

We use the Scottish CVD Policy Model for our cost-effectiveness analysis. The Scottish CVD Policy Model is a comprehensive simulation model developed to inform primary prevention and management strategies in Scotland. This model aims to quantify the long-term health impact and healthcare cost of various interventions targeted at reducing the burden of CVD, which is suitable for our target. The Scottish CVD Policy Model is an open-source, decision-analytic simulation model developed and validated using data from the Scottish population. Based on these ASSIGN risk profiles, which incorporates various individual-level risk factors, this model is able to project life expectancy, quality-adjusted life-years (QALYs), and healthcare costs for individuals receiving care within the Scottish National Health Service (NHS). The source code and programming files used to run the model are publicly accessible and can be downloaded at https://github.com/yiqiaoxin/CVDmodel.

Fig.1 shows a diagram of the state transition for this CVD Policy model. Individuals start in a CVD-free state and can transit to three distinct CVD event states plus a non-CVD death state. If a hospitalized patient died within 28 days of their admission, the first event was reclassified as fatal. Individuals who experience a non-fatal first CVD event can then transition to two additional non-fatal CVD event states before ultimately transitioning to the death state.

**Fig.1.**
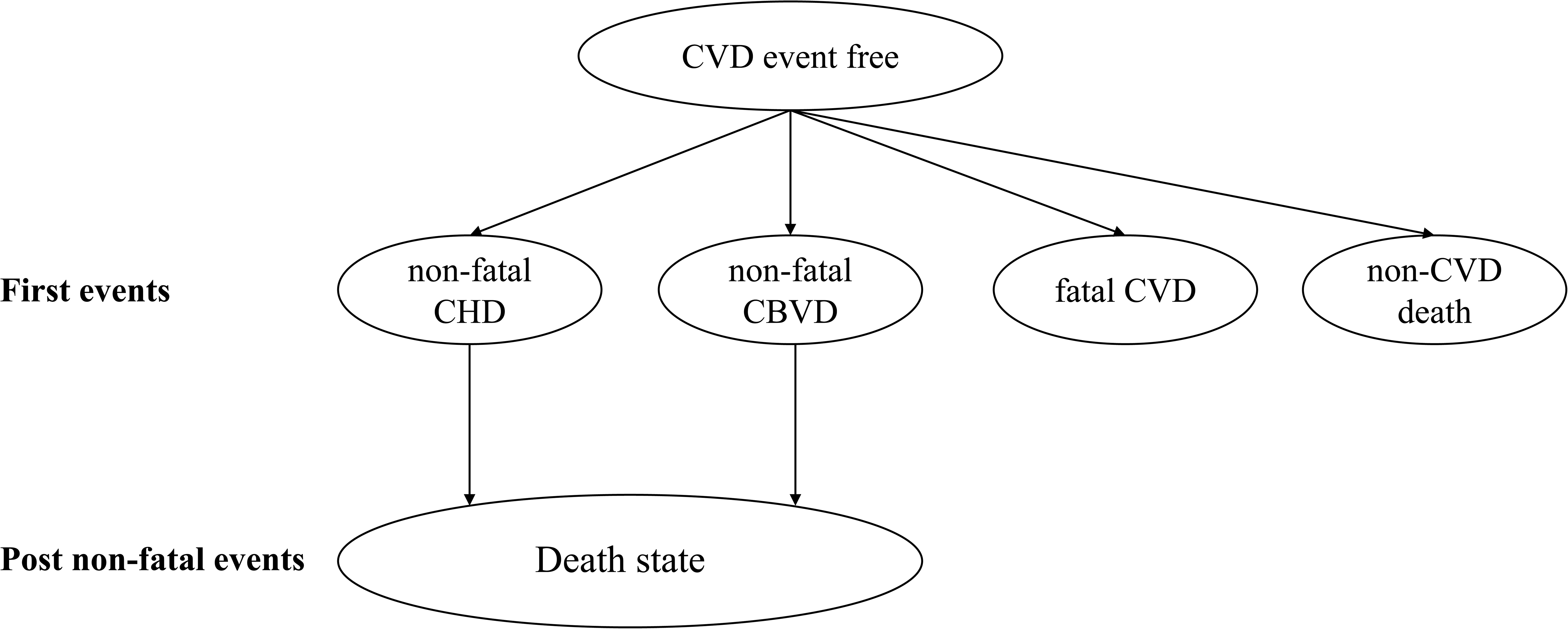
Structure of the Scottish Cerebrovascular Disease Policy Model. *CVD: cardiovascular disease; CHD: coronary heart disease; CBVD: cerebrovascular disease*.

The detailed process is outlined in the **appendix**.

#### 2.1.4 Intervention strategies

We compared the cost-effectiveness of statins and berberine therapies by analyzing different intervention strategies in the SHS cohort, and included nine scenarios, including no intervention, atorvastatin 20 mg per day (statins), berberine 1000 mg per day (berberine), simvastatin 20 mg plus berberine 1500 mg per day (combined intervention 1) and simvastatin 20 mg plus berberine 900 mg per day (combined intervention 2) for individuals with an ASSIGN score ≥20% (ASSIGN≥20%), and statins, berberine, combined intervention 1 and combined intervention 2 for individuals with an ASSIGN score ≥10% (ASSIGN≥10%). Given the limited RCT data on the combination of statins and berberine, which have been evaluated solely in mainland Chinese cohorts,[21] we selected two intervention strategies for simulation: simvastatin 20 mg plus berberine 1500 mg, and simvastatin 20 mg plus berberine 900 mg.[30, 31]

### 2.2 Model variables

The key parameter values used in the Scottish CVD Policy Model, including state transition probabilities, intervention effectiveness, utilities, and cost inputs, are presented in Table 1 and Appendix Tables 1, 2, 3, 4, 5, and 6.

**Table 1.** Intermediate-Intensity Intervention Measures of Statin, Berberine and Combined interventions.

| Parameters | Base case(95%CI) | Distribution | Data source | Reference |
| --- | --- | --- | --- | --- |
| Intervention effectiveness of statins |  |  |  |  |
| Non-HDL-c reduction, % |  |  |  |  |
| Atorvastatin 20mg | 26.0(15.0~35.0) | Beta | Meta-analysis | 10 |
| Berberine 1000mg | 16.4(14.6~18.2) | Beta | RCT | 36 |
| Simvastatin 20mg+ berberine 1500mg | 34.4(33.1~35.7) | Beta | RCT | 30 |
| Simvastatin 20mg+ berberine 900mg | 44.6(35.8~53.4) | Beta | RCT | 31 |
| HDL-c increase, % |  |  |  |  |
| Atorvastatin 20mg | 4.0(0.0~8.0) | Beta | Meta-analysis | 34,35 |
| Berberine 1000mg | 9.1(3.8~14.4) | Beta | RCT | 36 |
| Simvastatin 20mg plus berberine 1500mg | 25.0(23.2~26.8) | Beta | RCT | 30 |
| Simvastatin 20mg plus berberine 900mg | 17.4(8.6~26.2) | Beta | RCT | 31 |
| Relative risk per 1.0 mmol/L reduction in non-HDL-c |  |  |  |  |
| Nonfatal coronary heart disease | 0.77(0.75~0.80) | Beta | Meta-analysis | 10,35,37 |
| Nonfatal cerebrovascular disease | 0.87(0.83~0.91) | Beta | Meta-analysis | 10,35,37 |
| Fatal cardiovascular disease | 0.90(0.86~0.92) | Beta | Meta-analysis | 10,35,37 |
| Any cardiovascular disease | 0.79(0.77~0.81) | Beta | Meta-analysis | 10,35,37 |
| Side effects and intervention disutility |  |  |  |  |
| Type 2 diabetes absolute risk increase, % |  |  |  |  |
| Statins | 0.50(0.1~1.0) | Log-normal | Review | 35,38 |
| Berberine | -4.6(-6.1~-4.2) | Log-normal | Review&cohort | 38,40 |
| Combined statin and berberine | 0.0(-4.6~0.5) | Log-normal | Estimate | 38,39 |
| Annual pill-taking disutility | 0.002 (0.000~0.004) | Beta | Estimate | 35,41 |
| Annual treatment-related costs, £ |  |  |  |  |
| Intervention cost |  |  |  |  |
| Atorvastatin 20 mg/d | 13.44(8.99~23.99) | Gamma | Official tariff | 42 |
| Berberine 1000mg/d | 94.58(27.34~424.86) | Gamma | Estimate | 43 |
| Simvastatin 20mg plus berberine 1500mg/d | 150.86(50.00~646.28) | Gamma | Estimate | 42,43 |
| Simvastatin 20mg plus berberine 900mg/d | 103.57(33.59~391.36) | Gamma | Estimate | 42,43 |
| Monitoring(appointments and blood tests), first year | 102.51(63.51~141.51) | Gamma | Official tariff | 29,35,42,44 |
| Monitoring(appointments and blood tests), subsequent | 55.48(16.48~94.48) | Gamma | Official tariff | 29,35,42,44 |
| Other costs, £ |  |  |  |  |
| Risk assessment | 17.68(10.68~24.68) | Gamma | Official tariff | 29,35,42,44 |
| Annual type 2 diabetes treatment | 314.33(156.50~469.50) | Gamma | Official tariff | 29,35,42,44 |
| Other costing parameters |  |  |  |  |
| Annual discount rate, % | 3.5 | - | Estimate | 45 |

The state transition probabilities were derived through a competing risk parametric survival analysis, which integrated data from the SHHEC and the Scottish Morbidity Records (SMR) database.[32] The SMR data was collected from various healthcare settings, including hospitals and primary care, providing a detailed account of patient diagnoses, treatments, and outcomes. Each health state within the model is assigned a disutility value, which was derived from a survey of the Scottish population.[33] Individuals without first CVD events are attributed a baseline health-related quality of life, differentiated by age and sex. Those inhabiting the chronic non-fatal CVD states experience a decrement to this background utility, with the magnitude depending on the type of primary event. Furthermore, individuals in the chronic states may incur additional utility losses due to the occurrence of secondary CVD events. The healthcare costs associated with each health state were estimated using a combination of SMR data and national tariffs for elective and non-elective hospitalizations.

#### 2.2.1 Intervention effectiveness

Statins and berberine reduced CVD risk by lowering individuals’ non-high-density lipoprotein cholesterol (non–HDL-c) levels. Statins have been shown to produce a 26.0% reduction in non-HDL-c.[10, 34, 35] Similarly, berberine has been found to decrease non-HDL-c by approximate 16.4% in European population.[36] Furthermore, meta-analytic evidence suggests that these reductions in non-HDL-c is associated with relative risks of 0.77 for non-fatal coronary heart disease(CHD), 0.87 for non-fatal cerebrovascular disease (CBVD), and 0.90 for fatal CVD per 1.0 mmol/L decrease in non-HDL-c.[10] [37]

While evidence indicates that taking statins may increase the absolute risk of new-onset diabetes by 0.5%,[38] berberine has been shown to be effective in improving glucose metabolism and decreasing diabetes risk by about 4.6%.[39] [40] However, there is currently no evidence on the effects of the combined treatment of statins and berberine on diabetes, so we used the absolute risk of diabetes to be 0. Additionally, as previously,[41] an annual disutility of 0.002 QALYs was applied to account for the burden of daily pill-taking.

#### 2.2.2 Intervention costs

Statin costs were sourced from the British National Formulary,[42] and berberine costs were obtained from Amazon UK.[43] All individuals entering the model incurred screening costs. Regular checkups were assigned monitoring costs, which were largely derived from an analysis of unit costs for health and social care in England and Wales.[28, 44] Furthermore, additional costs were incorporated for each statin or berberine user to account for the differential risk of developing diabetes associated with these interventions.

### 2.3 Cost-effectiveness Analysis

The Scottish CVD Policy Model was employed to assess the cost-effectiveness of different drug intervention strategies for individuals with ASSIGN≥20%, as well as those with ASSIGN≥10%. First, we assessed the cost-effectiveness of stains, berberine and combined interventions in comparison to no intervention. Next, we assessed the cost-effectiveness of berberine and combined interventions relative to statin therapy. The cost-effectiveness analysis commenced with a baseline simulation to predict health and cost outcomes in the absence of any intervention. Subsequently, the model was adapted to incorporate the benefits, side effects, and costs associated with statin or berberine therapy. Individuals from the SHS cohort were simulated, and if they met the intervention criteria, they were assigned the corresponding outcomes. Otherwise, no intervention effects were applied.

The study adhered to the Consolidated Health Economic Evaluation Reporting Standards (CHEERS) guidelines. The primary outcome was the incremental cost per QALY gained for the different intervention strategies, considering a lifetime horizon. Intermediate outcomes, such as primary CVD events prevented, QALYs gained, and disaggregated costs, were also recorded. Future costs and health benefits were discounted at an annual rate of 3.5%.[45] A intervention strategy was deemed cost-effective if its incremental cost-effectiveness ratio (ICER) was less than £20,000 per QALY, a standard threshold used in cost-effectiveness analyses in Scotland and the United Kingdom.[46]

#### 2.3.1 Sensitivity analysis

The simulation analysis was conducted using a probabilistic approach. The parameter distributions and the risk factor hazard ratios in Table 1 were stochastically sampled. This allowed for the estimation of costs and QALYs for the respective intervention strategies in 1,000 independent iterations. The base-case results were derived from the mean values of the probabilistic analyses. Additionally, 95% confidence intervals were calculated as the 2.5th and 97.5th percentiles of the 1,000 iterations. Furthermore, cost-effectiveness acceptability curves were produced based on the results of the probabilistic analysis.

In the UK, statins are available through the National Health Service (NHS) as prescription medications, whereas berberine is classified as a nutritional supplement and is not covered, leading to significantly higher prices for berberine compared to statins.[42, 43, 47] This pricing structure may obscure the true cost-effectiveness of berberine. In contrast, in China, berberine is commonly used as a non-prescription medication, with a lower price that is comparable to that of statins.[48, 49] Therefore, to assess the potential impact of differing regional drug costs, we conducted a supplementary analysis using the prices of berberine and statins from China (Supplemental Table 1). We obtained the statins and berberine costs from Chinese sources and repeated the entire analysis to determine if the cost-effectiveness results would change compared to the primary analysis using UK-based drug prices.

## 3. Results

### 3.1 The CVD Risk Profile of the Final Intervention-Eligible Population

Supplemental Table 2 shows the CVD risk profile of the eligible population we included in the analysis. Women account for 55.6% of this population. On average, women had lower mean systolic blood pressure (SBP), higher mean TC level, and higher mean HDL-c level compared to men.

### 3.2 Cost-Effectiveness of Different Intervention Strategies

Table 2 and Fig.2 show the costs and QALY estimates for different intervention strategies in primary prevention, stratified by ASSIGN risk thresholds. Lowering the ASSIGN risk threshold from 20% to 10% increases the proportion of people in Scotland over 40 without CVD who are eligible for primary prevention intervention from 34.4% to 60.0%.

**Fig.2.**
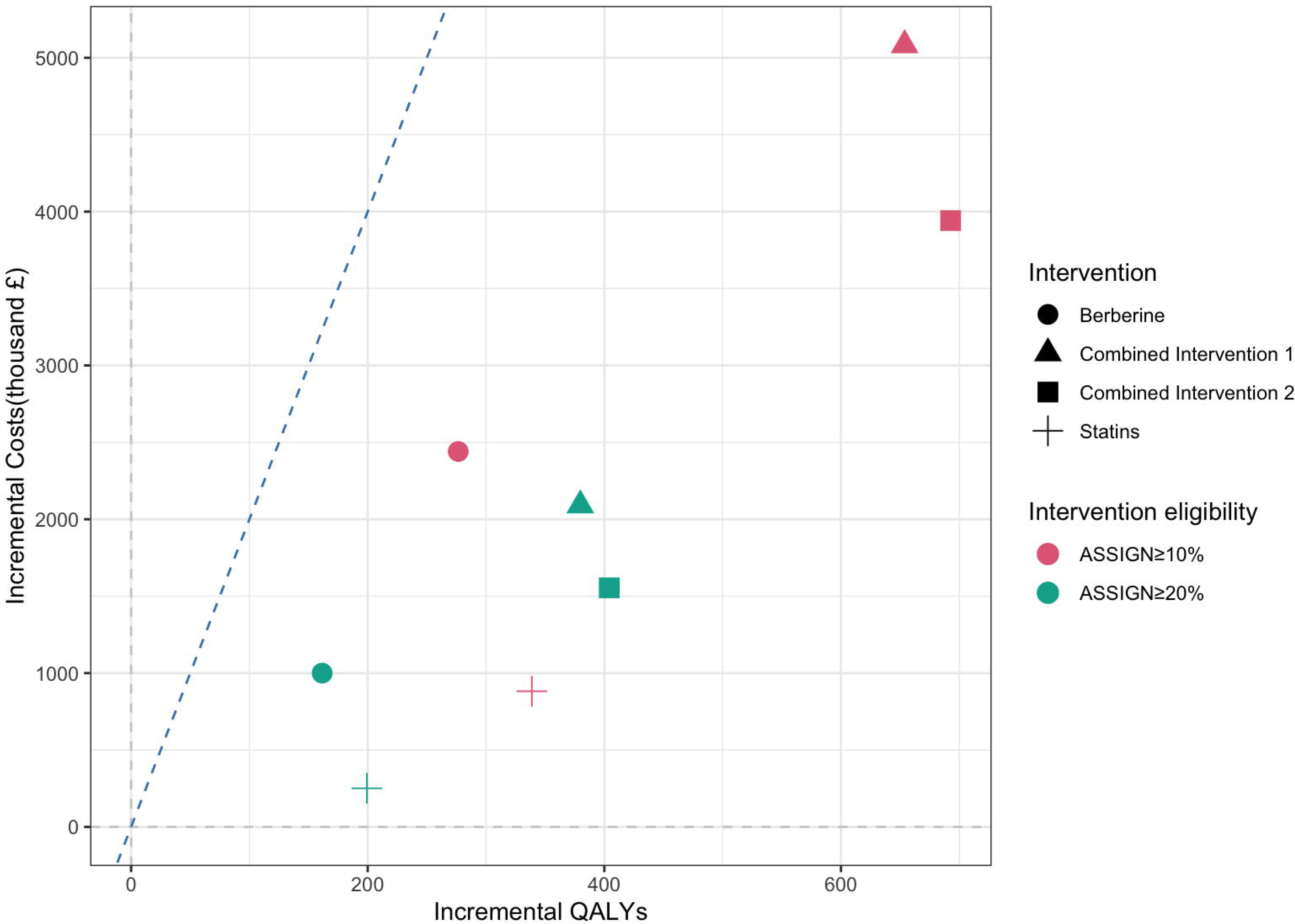
Cost-effectiveness plane for all intervention strategies. *The blue dashed line represents the cost-effectiveness threshold of £20,000/quality-adjusted life-year (QALY). An intervention strategy is considered cost-effective if the colorful point is below the blue dashed line*.

**Table 2.**
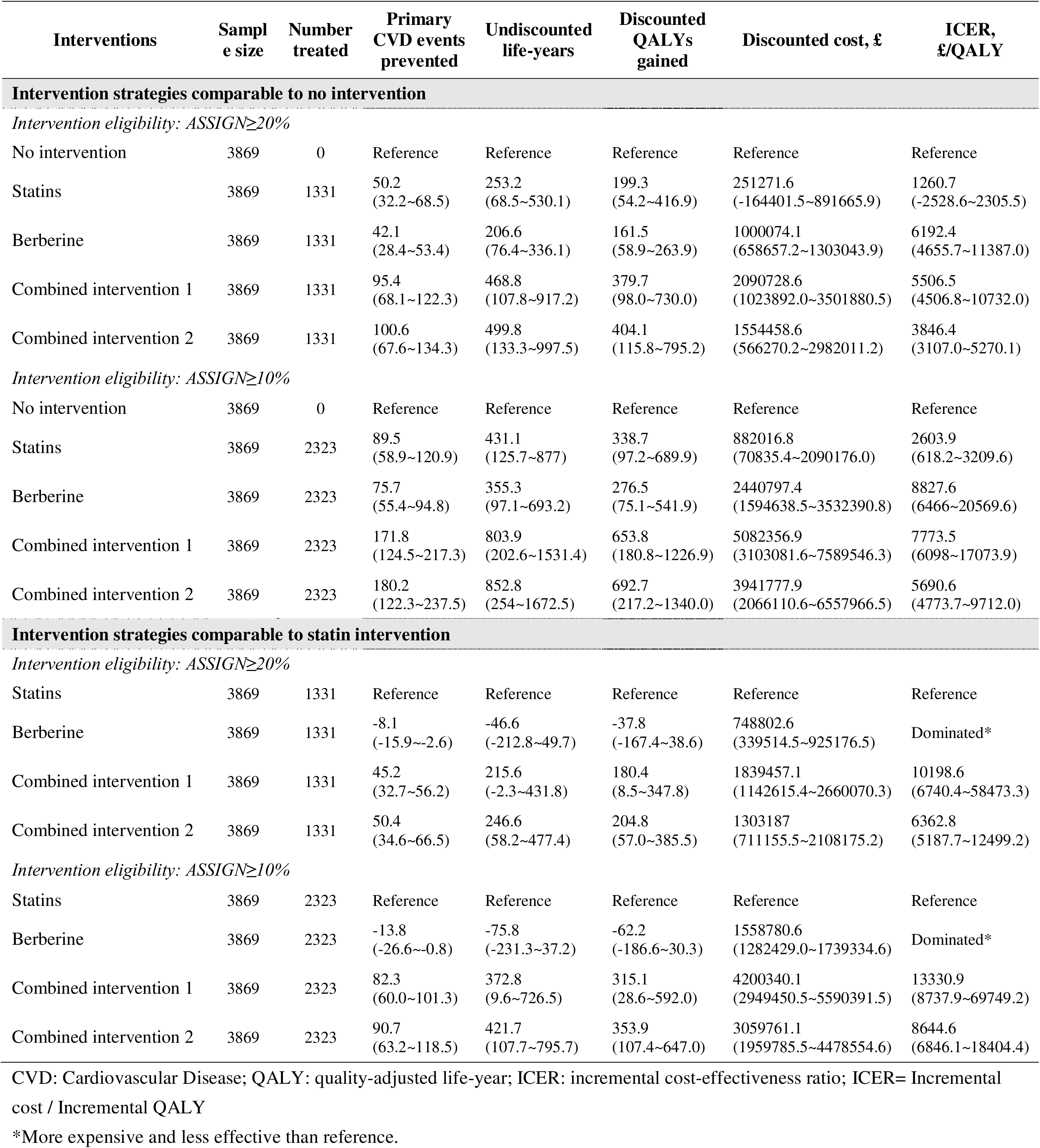
The cost-effectiveness of Statin, Berberine and Combined Interventions.

Compared to no intervention, at the threshold of £ 20,000 per QALY, all intervention strategies were found to be cost-effective in preventing primary CVD events. For individuals with ASSIGN≥20%, statin intervention was estimated to prevent 50.2 CVD events and produce an ICER of £1,260.7/QALY, berberine intervention was estimated to prevent 42.1 CVD events with an ICER of £6,192.4/QALY, combined intervention 1 was estimated to prevent 95.4 CVD events with an ICER of £5,506.5/QALY, and combined intervention 2 was estimated to prevent 100.6 CVD events with an ICER of £3,846.4/QALY. For individuals with ASSIGN≥10%, the corresponding estimates were 89.5 events (ICER £2,603.9/QALY) for statins, 75.7 events (ICER £8,827.6/QALY) for berberine, 171.8 events (ICER £7,773.5/QALY) for combined intervention 1, and 180.2 events (ICER £5,690.6/QALY) for combined intervention 2.

Compared to statin intervention, the berberine strategy was predicted to be more costly and less effective (i.e., dominated), but combined interventions remained cost-effective (combined intervention 1: ICER £10,198.6/QALY for ASSIGN≥20%, ICER £13,330.9/QALY for ASSIGN≥10%; combined intervention 2: ICER £6,362.8/QALY for ASSIGN≥20%, ICER £8,644.6/QALY for ASSIGN≥10%) at the threshold of £ 20,000 per QALY.

When using drug costs from China, where the price of berberine is reduced to below that of statins, all intervention strategies were still cost-effective compared to no intervention for both ASSIGN≥20% and ASSIGN≥10% thresholds. However, berberine alone or in conjunction with statins were also found to be cost-effective compared to statins alone for both ASSIGN risk levels. (Supplemental Table 1, 3, and Supplemental Figure 1)

### 3.3 Probability of Cost-Effectiveness

Probabilistic sensitivity analysis (Fig.3) showed that at cost-effectiveness thresholds below £20,000/QALY, all intervention strategies were cost-effective in most model iterations.

**Fig.3.**
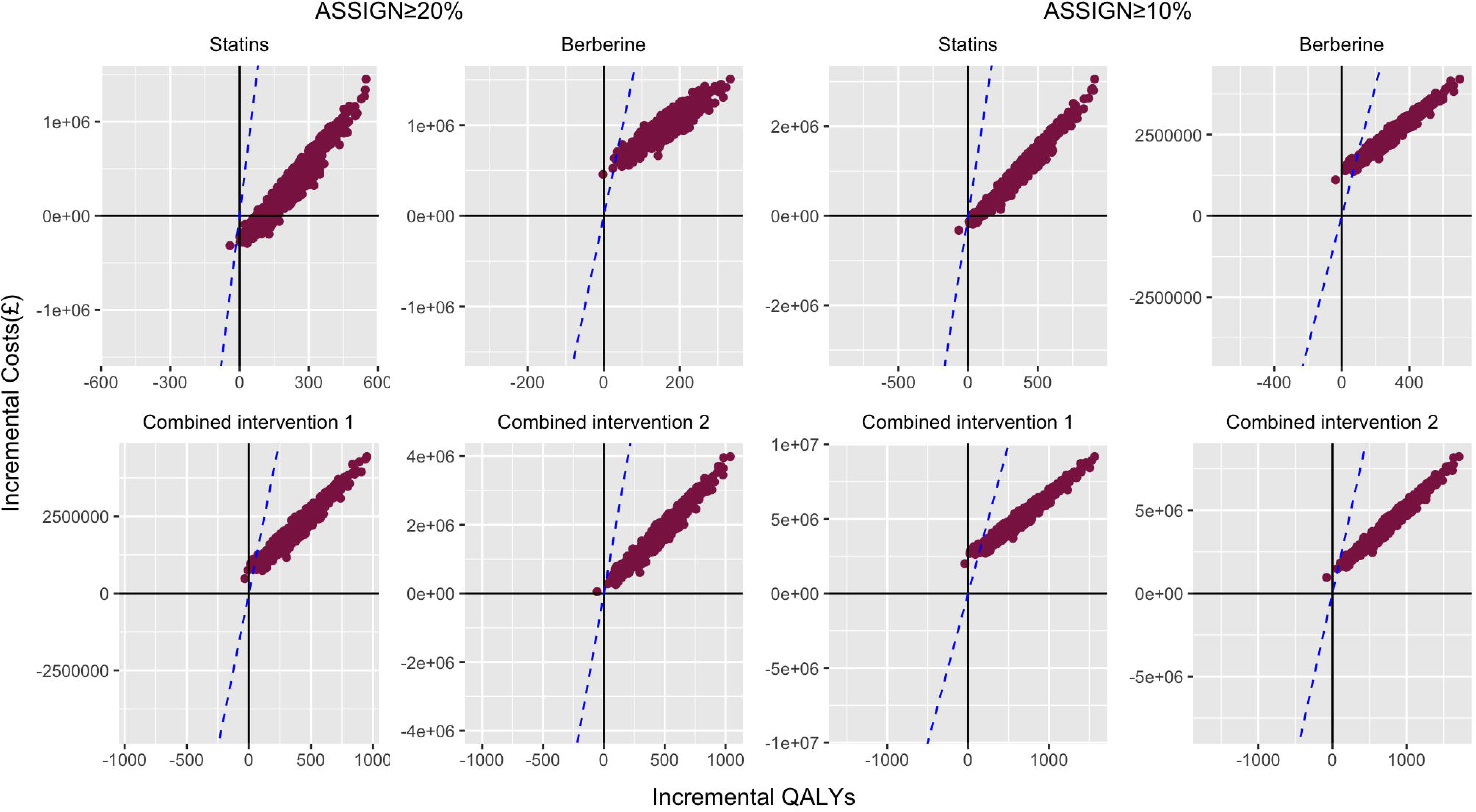
Probabilistic sensitivity analysis for all intervention strategies. *The blue dashed line represents the cost-effectiveness threshold of £20,000/quality-adjusted life-year (QALY). The proportions of dots below these blue dashed lines identify the likelihood that the Monte Carlo simulation results would yield an incremental cost-effectiveness ratio (ICER) below £20,000/QALY*.

Cost-effectiveness acceptability curves (Fig.4) indicated that at a £20,000/QALY threshold, statins, berberine, combined intervention 1, and combined intervention 2 were the optimal strategies 99.9%, 99.7%, 99.5%, and 99.9% of the time, respectively, for individuals with ASSIGN risk ≥20%. For those with ASSIGN risk ≥10%, the corresponding optimal strategy probabilities were 99.9%, 97.1%, 98.3%, and 99.8%.

**Fig.4.**
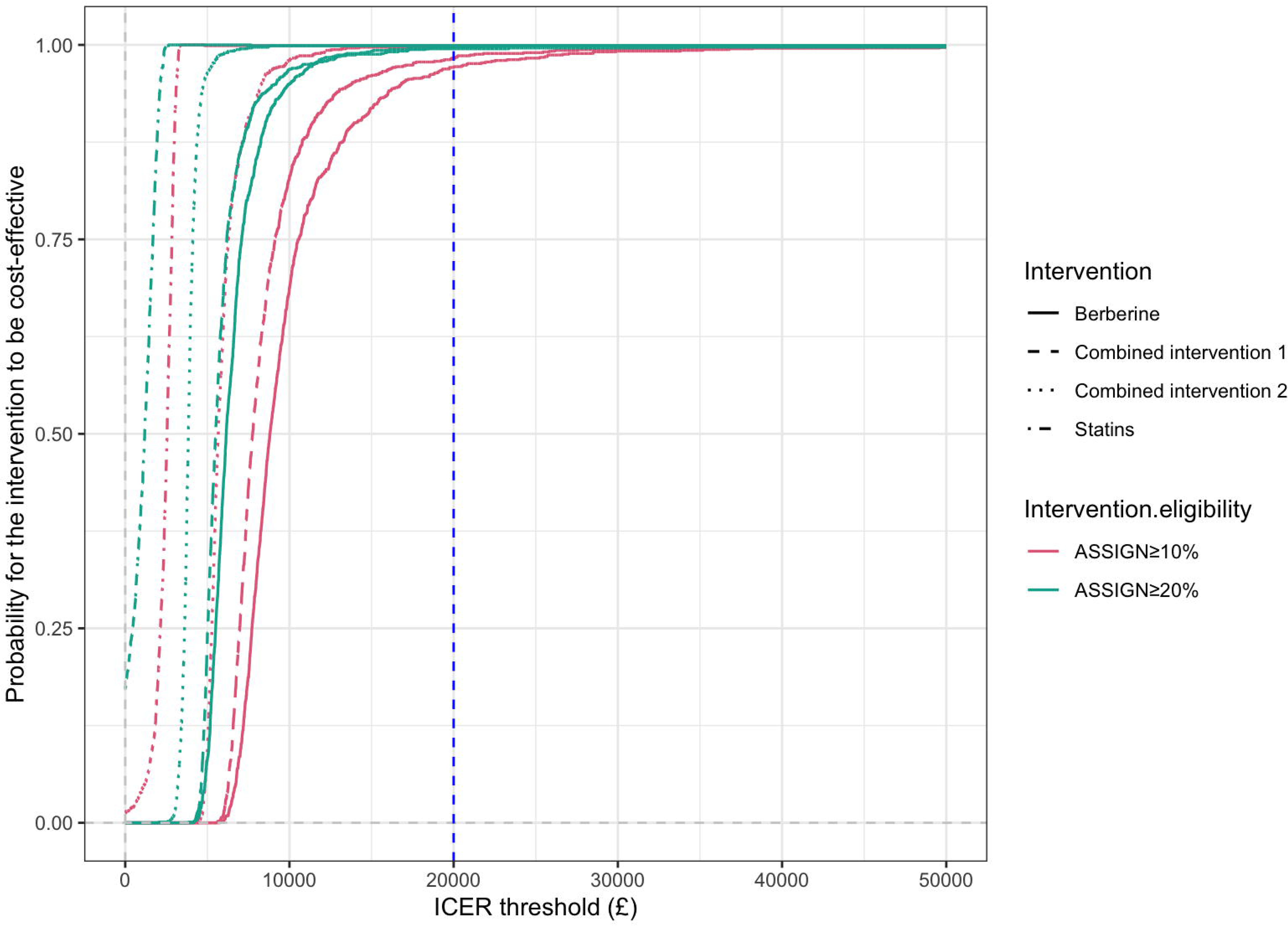
Cost-effectiveness acceptability curves for all intervention strategies. *The blue dashed line represents the cost-effectiveness threshold of £20,000/quality-adjusted life-year (QALY). The probabilities of statins, berberine, and combined interventions being cost-effective are over 95% at a £20,000/QALY threshold*.

The supplementary analysis, which assumed a reduced berberine price by switching to Chinese sources, yielded similar results (Supplemental Figure 2-3).

## 4. Discussion

This study provides a comprehensive cost-effectiveness analysis of various intervention strategies for the primary prevention of CVD. The analysis compared the cost-effectiveness of statin therapy, berberine supplementation, and combination approaches. The results indicate that compared to no intervention, statins, berberine, and combined interventions would be all cost-effective in preventing primary CVD events. When compared to statin intervention, berberine alone was less effective and more costly, and combined interventions remained cost-effective. However, if the price of berberine decreased to below that of statins, berberine alone or combined with statins was more cost-effective compared to statin monotherapy.

To our knowledge, this is the first comprehensive cost-effectiveness analysis examining berberine, both alone and in combination with statins, for primary CVD prevention. Previous studies have primarily focused on the cost-effectiveness of statins monotherapy or simplistic comparisons of lipid-lowering medications versus supplements.[35, 41, 50–53] This analysis provides a more robust assessment of the healthcare costs and QALYs associated with different intervention strategies. Compared to no intervention, statins, berberine, and combined interventions all demonstrate cost-effectiveness, indicating that high-risk populations have multiple options for the primary prevention of CVD events. For patients with hyperlipidemia who are intolerant to statins or seeking to optimize their lipid profiles through natural means, berberine offers an alternative as a nutraceutical, which can be used alone or in combination with statins. However, berberine is less cost-effective compared to statin intervention, while combined interventions remain cost-effective. This suggests that for patients with hyperlipidemia who tolerate statins, either statins alone or in combination with berberine is still the superior choice for primary prevention.

Our results confirm the cost-effectiveness of berberine but it is less cost-effective than statins alone at the cost in the setting of UK. Conversely, it would be more cost-effective in the setting where we used the cost in China. The UK’s healthcare system, primarily driven by the NHS, offers free medical services to residents and applies strict price controls on prescription medications, resulting in low statin prices.[47] However, berberine, categorized as a dietary supplement, is not covered by the NHS, and its price is determined by market supply and demand. In the UK, berberine needs to be imported from overseas, which increases costs related to transportation and tariffs. Additionally, the relatively low demand and limited competition contribute to its higher price. In contrast, in China, berberine is widely used in traditional Chinese medicine, leading to high market demand and intense competition.[54, 55] There are numerous berberine cultivation bases and production factories, resulting in lower production and transportation costs, and consequently, lower prices. If the UK were to include berberine in the NHS coverage or reduce its price to a moderate level, the strategies of using berberine alone or in combination with statins would be more cost-effective, similar to findings based on Chinese pricing. This would provide an alternative for individuals with statin intolerance or serious side effects, enabling them to pursue primary prevention.

We expect that the use of berberine, either alone or in combination with statins, will enhance the clinical guidelines and policies for primary CVD prevention. In 2018, the International Lipid Expert Panel (ILEP) suggested considering berberine for lipid-lowering intervention in patients who are intolerant to statins; however, the evidence at that time was limited.[24] The 2019 ESC/EAS guidelines on the management of dyslipidemia also acknowledged the lipid-lowering effects of dietary supplements and functional foods, including berberine.[25] Our findings demonstrate that berberine offers greater cost-effectiveness compared to statins when its price is moderate. This provides additional evidence for the use of berberine in patients intolerant to statins and supports the refinement of clinical guidelines and policies for CVD prevention.

This study has several notable strengths. First, the primary outcome was the occurrence of first CVD events or deaths, which provides a clinically relevant endpoint. However, the analysis also modeled the QALYs and costs throughout lifetime, accounting for potential side effects of medications such as diabetes. This comprehensive approach offers a more complete picture of the cost-effectiveness of primary CVD prevention strategies. Additionally, the analysis considered the context-specific pricing of berberine. In the UK, berberine is priced as a high-cost nutritional supplement.[43] In contrast, berberine is widely used as a lower-cost drug in China due to its proven effectiveness.[21] Leveraging the Chinese pricing data can provide valuable supplementary support for evaluating the health economic viability of berberine-based interventions. If other countries are able to similarly reduce the costs of berberine, it could further enhance its cost-effectiveness as a primary prevention option.

However, the analysis also has several limitations. When estimating the cost-effectiveness of combined intervention, the model parameters were derived from RCTs conducted in Chinese populations.[30, 31] The sample sizes of these RCTs were relatively small, and the overall quality of the evidence was not optimal, which introduces potential bias into the final results. Currently, there is a paucity of high-quality RCT data evaluating the efficacy and safety of combined therapy approaches for managing hyperlipidemia and preventing CVD outcomes. The available evidence is primarily from the Chinese context. Despite these limitations, this study offers a valuable contribution by comprehensively modeling the lifetime cost-effectiveness of different intervention strategies, including combined interventions, to inform evidence-based decision-making for primary CVD prevention.

## 5. Conclusion

This comprehensive cost-effectiveness analysis found that statins, berberine, and combined interventions were all cost-effective strategies for primary CVD prevention. Compared to statin intervention, berberine alone was less cost-effective, while the combined statin-berberine approach remained cost-effective. However, a reduction in the price of berberine could render berberine alone or in combination with statins as cost-effective alternatives. This study provides valuable insights to guide evidence-based decision-making and resource allocation for primary CVD prevention. The findings highlight the potential role of berberine as an alternative or complementary intervention option, particularly if its price decreases to below that of statins.

## Supporting information

Apendix

## Statements and Declarations

### Ethics approval and consent to participate

Original ethical approval for the Scottish Health Survey series was granted by the Research Committee for Wales and participants gave full informed consent to participate in the study. Anonymised secondary data are accessible via the UK Data Service, for which no additional ethical approval was required.

## Data availability statement

Available from: DOI: http://doi.org/10.5255/UKDA-Series-2000047

## Acknowledgments

We would like to acknowledge that the Scottish Health Survey provided the data used for this study through the UK Data Service. Available from: DOI: http://doi.org/10.5255/UKDA-Series-2000047

## Authors’ contributions

Conceptualization: Jie V. Zhao; Methodology: Jie V. Zhao, Kathy Leung and Yuanqing Xia; Formal analysis and investigation: Yuanqing Xia; Writing - original draft preparation: Yuanqing Xia; Writing - review and editing: Jie V. Zhao, Kathy Leung and Yuanqing Xia; Funding acquisition: Jie V. Zhao and Kathy Leung; Supervision: Jie V. Zhao and Kathy Leung.

## Abbreviations List

CVD: Cardiovascular Disease
CHD: coronary heart disease
CBVD: cerebrovascular disease
SIMD: Scottish Index of Multiple Deprivation
SBP: Systolic Blood Pressure
TC: Total Cholesterol
HDL-c: High-Density Lipoprotein Cholesterol
LDL-c: Low-Density Lipoprotein Cholesterol
non–HDL-c: non-high-density lipoprotein cholesterol
QALY: quality-adjusted life-year
ICER: incremental cost-effectiveness ratio
RCT: randomized clinical trial

## Supplementary legends

**Supplemental Table 1** Annual statin and berberine costs from Chinese source.

**Supplemental Table 2** Comparison of CVD risk factors for the final intervention-eligible population.

**Supplemental Table 3** The cost-effectiveness of Statin, Berberine, and Combined Interventions in supplementary analyses.

**Supplemental Figure 1** Cost-effectiveness plane for all intervention strategies in supplementary analyses.

*The blue dashed line represents the cost-effectiveness threshold of £20,000/quality-adjusted life-year (QALY). An intervention strategy is considered cost-effective if the colorful point is below the blue dashed line.*

**Supplemental Figure 2** Probabilistic sensitivity analysis for all intervention strategies in supplementary analyses.

*The blue dashed line represents the cost-effectiveness threshold of £20,000/quality-adjusted life-year (QALY). The proportions of dots below these blue dashed lines identify the likelihood that the Monte Carlo simulation results would yield an incremental cost-effectiveness ratio (ICER) below £20,000/QALY*.

**Supplemental Figure 3** Cost-effectiveness acceptability curves for all intervention strategies in supplementary analyses.

*The blue dashed line represents the cost-effectiveness threshold of £20,000/quality-adjusted life-year (QALY). The probabilities of statins, berberine, and combined interventions being cost-effective are over 99% at a £20,000/QALY threshold*.

