## Supplementary material for "Cost-Effectiveness Analysis of Statins, Berberine, and Their Combined Use for Primary Prevention of Cardiovascular Disease": Apendix

### Appendix

The Scottish CVD Policy Model

This cardiovascular disease (CVD) policy model is intended to inform primary prevention policy to avoid premature morbidity and mortality and associated health service costs, which is suitable for our target.[32,33] **Figure 1** shows a diagram of the model. Individuals enter the model CVD-free and subsequently transition to one of four primary event states: non-fatal coronary heart disease (CHD), non-fatal cerebrovascular disease (CBVD), fatal CVD, or fatal non-CVD, until death state.

1. State Transition Probabilities

The model's state transitions are of two types: 1) transitions to primary events (fatal or non-fatal), and 2) transitions to all-cause mortality after a non-fatal primary event. These transition probabilities were derived from competing risks survival analysis of linked Scottish health data.

Specifically, the model parameters were estimated using the Scottish Heart Health Extended Cohort (SHHEC) and Scottish Morbidity Records (SMR) datasets.[35,56] SHHEC provided baseline risk factor information on age, sex, total cholesterol (TC), high-density lipoprotein cholesterol (HDL-c), systolic blood pressure (SBP), family history of CVD, diabetes, cigarettes smoked per day (CPD), and Scottish Index of Multiple Deprivation (SIMD). These will be referred to hereafter as the ASSIGN risk factors. The SMR database captured subsequent hospitalization events. Linking these datasets allowed researchers to analyze the relationship between individual characteristics and cardiovascular outcomes.

A parametric competing risks approach was used to estimate cause-specific hazard functions for the four primary events: non-fatal CHD, non-fatal CBVD, fatal CVD, and fatal non-CVD. Gompertz regression models were fit with the ASSIGN risk factors as covariates. Results for primary event regression, developed by Lewsey et al., are presented in **Appendix Table 1**.[32]

Transition probabilities from non-fatal events to all-cause mortality were similarly estimated using Gompertz regression. In this regression, age at first event, SIMD and family history of CVD were employed as covariates. Results for the secondary event regressions, also developed by Lewsey et al., are presented in **Appendix Table 2**.[32]

Additionally, the model accounts for the occurrence of secondary health events (e.g. angina, myocardial infarction) within the chronic disease states, with probabilities determined through probit regression on age at first event, SIMD, and family history of CVD (**Appendix Table 3**).[33]

2. Health-Related Quality of Life

Health-related quality of life (HRQoL) inputs for the model were derived from the 2003 Scottish Health Survey (SHS), the most recent large-scale survey of the Scottish population to measure quality of life. A total of 7,054 respondents aged 20 and older completed the 12-item Short Form (SF-12) HRQoL questionnaire, which was used to generate QALY values. **Appendix Table 4** shows the mean HRQoL scores by age and socioeconomic group from the SHS 2003 dataset. These scores are used to weight survival probabilities across the model arms.

Linear regression was performed on the SHS 2003 data to estimate baseline QALY values for the Scottish adult population and to quantify utility decrements associated with various cardiovascular events. The dependent variable was the SF-12-derived QALY score, with independent variables including sex, age, and six CVD events. These regression results are presented in **Appendix Table 5**.

3. Healthcare Costs

Lifetime hospitalization costs were estimated for all individuals in the SHHEC-SMR dataset as a function of the events experienced and the corresponding length of stays. The costed SHHEC-SMR dataset was then analyzed to predict health state-related costs in the model.

The approach outlined by Geue et al. was used to attribute costs to each hospitalization episode in the dataset.[57] This involved assigning a healthcare resource group (HRG) to each episode using HRGv3.5 Grouper software, followed by applying the English NHS tariff. To avoid overestimating costs for continuous inpatient stays (CIS) involving multiple episodes, a ‘Spell Converter’ software was employed to designate a dominant episode for each CIS.

The estimated lifetime hospitalization costs for each individual were then used in linear regression models to predict pre- and post-event hospitalization costs for the model (**Appendix Table 6**). Cubic splines were included to capture non-linearity over time, and other covariates such as age at model entry (for pre-event costs), age at first event (for post-event costs), SIMD, and family history of CVD.

Appendix Table 1. Cause-specific hazards of primary first events

| **Covariate** | **first_event_nonfatal_CHD** | **first_event_nonfatal_CBVD** | **first_event_CVD_death** | **first_event_nonCVD_death** |
| --- | --- | --- | --- | --- |
| **Male** |  |  |  |  |
| age | 1.046(1.039-1.053) | 1.068(1.055-1.081) | 1.097(1.085-1.108) | 1.099(1.089-1.107) |
| SIMD | 1.004(1.001-1.007) | 1.009(1.005-1.014) | 1.006(1.003-1.01) | 1.009(1.007-1.012) |
| diabetes | 1.921(1.339-2.757) | 3.216(1.943-5.328) | 2.37(1.476-3.808) | 1.398(0.845-2.314) |
| Family History of CVD | 1.504(1.343-1.685) | 0.979(0.791-1.213) | 1.179(0.997-1.394) | 0.985(0.85-1.142) |
| No. of cigarette per day | 1.018(1.013-1.022) | 1.024(1.017-1.031) | 1.031(1.026-1.038) | 1.031(1.026-1.036) |
| Systolic Blood Pressure | 1.008(1.005-1.011) | 1.012(1.007-1.016) | 1.015(1.012-1.019) | 0.999(0.995-1.002) |
| TC | 1.29(1.231-1.353) | 1.087(0.998-1.183) | 1.127(1.051-1.208) | 0.95(0.895-1.008) |
| HDL | 0.468(0.388-0.563) | 0.882(0.674-1.155) | 0.867(0.691-1.089) | 1.468(1.239-1.74) |
| **Female** |  |  |  |  |
| age | 1.06(1.05-1.069) | 1.083(1.067-1.1) | 1.107(1.091-1.123) | 1.095(1.084-1.106) |
| SIMD | 1.009(1.006-1.012) | 1.013(1.009-1.018) | 1.004(1-1.009) | 1.007(1.004-1.01) |
| diabetes | 2.065(1.409-3.028) | 3.007(1.813-4.988) | 3.139(1.974-4.998) | 0.964(0.513-1.811) |
| Family History of CVD | 1.675(1.476-1.902) | 1.428(1.162-1.754) | 1.27(1.051-1.534) | 0.982(0.848-1.138) |
| No. of cigarette per day | 1.021(1.014-1.027) | 1.027(1.017-1.038) | 1.049(1.041-1.058) | 1.039(1.033-1.045) |
| Systolic Blood Pressure | 1.006(1.003-1.009) | 1.014(1.009-1.018) | 1.018(1.013-1.022) | 1.003(0.999-1.006) |
| TC | 1.207(1.146-1.271) | 0.95(0.861-1.049) | 1.059(0.978-1.146) | 0.927(0.869-0.987) |
| HDL | 0.474(0.392-0.574) | 0.708(0.529-0.946) | 0.84(0.657-1.076) | 0.956(0.795-1.148) |

CVD: Cardiovascular Disease; CHD: coronary heart disease; CBVD: cerebrovascular disease; SIMD: Scottish Index of Multiple Deprivation; TC: Total Cholesterol; HDL: High-Density Lipoprotein Cholesterol

Appendix Table 2. Cause-specific hazards of post-CVD mortality

| **Covariate** | **post_CHD** | **post_CBVD** |
| --- | --- | --- |
| **Male** |  |  |
| Age at event | 1.08(1.067-1.094) | 1.069(1.049-1.091) |
| SIMD score | 1.013(1.009-1.017) | 1.009(1.003-1.015) |
| Family history | 0.966(0.792-1.176) | 1.063(0.769-1.47) |
| **Female** |  |  |
| Age at event | 1.077(1.061-1.093) | 1.073(1.051-1.093) |
| SIMD score | 1.007(1.003-1.012) | 1(0.993-1.008) |
| Family history | 0.752(0.595-0.95) | 1.198(0.859-1.67) |

CVD: Cardiovascular Disease; CHD: coronary heart disease; CBVD: cerebrovascular disease; SIMD: Scottish Index of Multiple Deprivation

Appendix Table 3. Probit regression, probability of various secondary non-fatal CVD events within chronic disease states

| **Covariate** | **Probit regression coefficient(95%CI)** | | | | |
| --- | --- | --- | --- | --- | --- |
|  | **CHD** | **Stroke** | **Intermittent Claudication** | **Other Heart Condition** | **Heart Failure** |
| After first non-fatal CHD | |  |  |  |  |
| **Male** |  |  |  |  |  |
| t1 | -0.019 (-0.040-0.002) | -0.101 (-0.147--0.055) | -0.049 (-0.104-0.006) | -0.06 (-0.091--0.028) | -0.152 (-0.195--0.11) |
| t2 | 0.071 (0.045-0.098) | 0.098 (0.035-0.161) | 0.034 (-0.039-0.107) | 0.05 (0.008-0.092) | 0.16 (0.098-0.223) |
| Age at first event (years) | 0.010 (0.005-0.016) | 0.001 (-0.007-0.009) | -0.001 (-0.012-0.010) | 0.005 (-0.002-0.013) | 0.014 (0.004-0.023) |
| SIMD | 0.003 (0.001-0.005) | 0.003 (0.000-0.007) | 0.002 (-0.003-0.006) | 0.003 (0.001-0.006) | 0.002 (-0.002-0.006) |
| Family history | 0.106 (0.019-0.193) | -0.011 (-0.168-0.145) | -0.136 (-0.350-0.077) | 0.174 (0.051-0.297) | 0.044 (-0.121-0.21) |
| Constant | -2.012 (-2.420--1.605) | -2.085 (-2.640--1.530) | -2.137 (-2.903--1.371) | -2.187 (-2.68--1.693) | -2.626 (-3.26--1.992) |
| **Female** |  |  |  |  |  |
| t1 | -0.003 (-0.033-0.027) | -0.072 (-0.144-0.000) | -0.078 (-0.152--0.004) | -0.045 (-0.087--0.003) | -0.119 (-0.176--0.061) |
| t2 | 0.057 (0.011-0.10 | 0.026 (-0.102-0.155) | 0.106 (-0.015-0.228) | 0.026 (-0.045-0.096) | 0.133 (0.039-0.227) |
| Age at first event (years) | 0.010 (0.004-0.016) | 0.007 (-0.006-0.019) | 0.016 (-0.001-0.033) | 0.012 (0.003-0.021) | 0.018 (0.000-0.029) |
| SIMD | 0.001 (-0.001-0.004) | 0.005 (0.001-0.009) | 0.001 (-0.005-0.008) | 0.001 (-0.002-0.004) | 0.004 (-0.000-0.009) |
| Family history | 0.056 (-0.046-0.158) | -0.014 (-0.221-0.194) | -0.129 (-0.379-0.122) | -0.138 (-0.27--0.005) | -0.038 (-0.246-0.17) |
| Constant | -2.137 (-2.58--1.694) | -2.638 (-3.492--1.783) | -3.274 (-4.558--1.990) | -2.407 (-3.00--1.81) | -3.017 (-3.587--2.177) |
| After first non-fatal CBVD | |  |  |  |  |
| **Male** |  |  |  |  |  |
| t1 | -0.069 (-0.150-0.012) | -0.035 (-0.087-0.017) | 0.020 (-0.085-0.125) | -0.070 (-0.134--0.006) | -0.129 (-0.275-0.017) |
| t2 | 0.063 (0.049--0.174) | 0.046 (-0.030-0.123) | -0.108 (-0.305-0.09) | 0.077 (-0.016-0.17) | 0.159 (-0.021-0.340) |
| Age at first event (years) | -0.003 (-0.016-0.010) | 0.010 (-0.001-0.021) | 0.001 (-0.016-0.019) | 0.004 (-0.009-0.017) | 0.039 (0.009-0.070) |
| SIMD | -0.002 (-0.008-0.004) | 0.003 (0.000-0.006) | 0.007 (0.001-0.014) | 0.002 (-0.002-0.007) | -0.010 (-0.021-0.001) |
| Family history | 0.144 (-0.085-0.373) | 0.019 (-0.154-0.191) | 0.024 (-0.34-0.389) | 0.053 (0.169-0.275) | 0.353 (-0.063-0.77) |
| Constant | -1.506 (-2.42--1.605) | -2.109 (-2.891--1.327) | -2.744 (-4.034--1.454) | -1.923 (-2.789--1.058) | -4.697 (-6.983--2.41) |
| **Female** |  |  |  |  |  |
| t1 | 0.078 (-0.028-0.183) | -0.023 (-0.077-0.03) | 0.008 (-0.137-0.152) | -0.053 (-0.138-0.032) | -0.186 (-0.319--0.054) |
| t2 | -0.088 (-0.225-0.049) | 0.056 (-0.02-0.132) | -0.069 (-0.275-0.137) | 0.034 (-0.071-0.140) | 0.227 (0.069-0.386) |
| Age at first event (years) | -0.000 (-0.014-0.013) | 0.022 (0.013-0.030) | -0.011 (-0.031-0.009) | 0.006 (-0.005-0.017) | -0.002 (-0.017-0.014) |
| SIMD | 0.004 (-0.003-0.011) | 0.001 (-0.002-0.004) | 0.000 (-0.013-0.014) | -0.001 (-0.006-0.004) | 0.001 (-0.007-0.010) |
| Family history | 0.068 (-0.222-0.359) | 0.042 (-0.192-0.108) | -0.303 (-0.798-0.192) | 0.281 (0.044-0.517) | 0.036 (-0.36-0.432) |
| Constant | -2.531 (-3.653--1.409) | -2.777 (-3.424--2.130) | -1.714 (-3.543-0.115) | -2.178 (-2.983--1.373) | -1.881 (-3.219--0.542 |

CVD: Cardiovascular Disease; CHD: coronary heart disease; CBVD: cerebrovascular disease; t1: time spline 1; t2: time spline 2; SIMD: Scottish Index of Multiple Deprivation

Appendix Table 4. Mean (and 95% CI) HRQoL scores in the general Scottish population by age group and fifths of SIMD groups

| **Age group (years)** | **SIMD 1 (least deprived)** | **SIMD 2** | **SIMD 3** | **SIMD 4** | **SIMD 5 (most deprived)** |
| --- | --- | --- | --- | --- | --- |
| Male |  |  |  |  |  |
| 35-44 | 0.834(0.816, 0.852) | 0.838(0.820, 0.855) | 0.823(0.805, 0.841) | 0.811(0.787, 0.835) | 0.777(0.744, 0.810) |
| 45-54 | 0.825(0.805, 0.845) | 0.827(0.806, 0.849) | 0.808(0.783, 0.833) | 0.791(0.756, 0.826) | 0.762(0.729, 0.794) |
| 55-64 | 0.845(0.826, 0.865) | 0.803(0.780, 0.826) | 0.820(0.796, 0.843) | 0.782(0.751, 0.813) | 0.718(0.680, 0.757) |
| 65-74 | 0.813(0.784, 0.841) | 0.822(0.791, 0.853) | 0.802(0.775, 0.830) | 0.761(0.729, 0.792) | 0.732(0.697, 0.768) |
| 75+ | 0.797(0.750, 0.843) | 0.802(0.770, 0.835) | 0.756(0.722, 0.791) | 0.775(0.731, 0.818) | 0.732(0.685, 0.779) |
| Female |  |  |  |  |  |
| 35-44 | 0.837(0.823, 0.852) | 0.827(0.812, 0.841) | 0.794(0.773, 0.815) | 0.788(0.769, 0.808) | 0.748(0.722, 0.774) |
| 45-54 | 0.827(0.812, 0.843) | 0.793(0.773, 0.812) | 0.780(0.758, 0.802) | 0.769(0.745, 0.792) | 0.736(0.708, 0.764) |
| 55-64 | 0.835(0.816, 0.854) | 0.815(0.798, 0.832) | 0.791(0.768, 0.814) | 0.769(0.742, 0.796) | 0.701(0.670, 0.732) |
| 65-74 | 0.827(0.803, 0.851) | 0.803(0.776, 0.830) | 0.792(0.766, 0.818) | 0.742(0.709, 0.776) | 0.702(0.668, 0.736) |
| 75+ | 0.741(0.702, 0.779) | 0.765(0.732, 0.798) | 0.715(0.681, 0.748) | 0.693(0.655, 0.731) | 0.689(0.652, 0.726) |

SIMD: Scottish Index of Multiple Deprivation

Appendix Table 5. Event utility decrements

| **Covariate** | **Utility decrements (95%CI)** |
| --- | --- |
| Male |  |
| CHD | 0.043 (0.019, 0.068) |
| Stroke | 0.092 (0.061, 0.122) |
| Intermittent claudication | 0.025 (-0.005, 0.056) |
| Other heart condition | 0.043 (0.011, 0.074) |
| Female |  |
| CHD | 0.037 (0.007, 0.067) |
| Stroke | 0.097 (0.067, 0.127) |
| Intermittent claudication | 0.017 (-0.009, 0.043) |
| Other heart condition | 0.023 (-0.011, 0.058) |

CHD: coronary heart disease

Appendix Table 6. Linear regression coefficient(95%CI), costs pre- and post-first events

| **Covariate** | **Pre-first-Event Costs, Linear Regression Coefficient(95%CI)** | | | | **Post-first-Event Costs, Linear Regression Coefficient(95%CI)** | |
| --- | --- | --- | --- | --- | --- | --- |
|  | **Non-fatal CHD** | **Non-fatal CBVD** | **Fatal CVD** | **Fatal non-CVD** | **Post non-fatal CHD** | **Post non-fatal CBVD** |
| **Male** |  |  |  |  |  |  |
| t1 | 18.6 (-2.0, 39.2) | 5.5 (-38.0, 49.0) | 26.1 (-7.1, 59.3) | 42.1 (-4.5, 88.7) | -552.6 (-638.7, -466.6) | -680.0 (-854.7, -505.2) |
| t2 | 115.0 (70.3, 159.8) | 156.6 (72.0, 241.2) | 114.6 (56.8, 172.5) | 237.2 (157.1, 317.4) | 654.9 (554.5, 755.3) | 787.7 (555.7, 1020) |
| Age at first event (years) | 22.7 (16.4, 29.0) | 17.3 (5.8, 28.9) | 27.7 (16.5, 38.89) | 24.7 (9.0, 40.5) | 84.6 (66.9, 102.4) | 112.6 (81.2, 144.0) |
| SIMD | 5.2 (3.0, 7.4) | 6.6 (2.9, 10.3) | 3.8 (-0.1, 7.8) | 4.7 (-0.5, 10.0) | 14.2 (7.6, 20.8) | 6.8 (-4.5, 18.1) |
| Family history | 93.8 (1.5, 186.1) | -161.9 (-328.5, 4.8) | 67.4 (-120.9, 255.7) | 116.9 (-198.7, 432.6) | 239.8 (-80.4, 560.0) | -102.2 (-717.2, 512.9) |
| Constant | -1121 (-1446, -795) | -832.8 (-1483, -182.4) | -1345 (-1981, -709.2) | -1029 (-1890, -169.2) | -1024 (-2107, 59.0) | -1836 (-4010, 338.3) |
| **Female** |  |  |  |  |  |  |
| t1 | -7.3 (-94.3, 79.8) | 14.2 (-23.3, 51.6) | 23.9 (-18.4, 66.2) | 59.0 (11.5, 106.4) | -548.6 (-652.4, -444.8) | -542.3 (-744.1, -340.4) |
| t2 | 172.2 (-64.6, 408.9) | 121.8 (57.7, 185.8) | 144.7 (72.2, 217.3) | 202.6 (125.8, 279.4) | 745.4 (600.4, 890.3) | 595.6 (357.4, 833.9) |
| Age at first event (years) | 0.5 (-25.1, 26.2) | 26.3 (15.0, 37.6) | 33.7 (18.4, 49.0) | 16.0 (-3.5, 35.4) | 90.7 (68.5, 112.9) | 97.1 (67.0, 127.2) |
| SIMD | 10.6 (2.3, 19.0) | 8.4 (2.4, 14.4) | 5.5 (0.8, 10.2) | 11.9 (5.6, 18.3) | 13.6 (6.0, 21.3) | 7.7 (-4.7, 20.0) |
| Family history | 337.8 (-235.7, 911.4) | 22.7 (-176.6, 222.0) | 105.5 (-125.0, 336.0) | 47.7 (-229.7, 325.2) | -227.9 (-596.5, 140.7) | -93.9 (-656.1, 468.4) |
| Constant | -214.2 (-1359, 930.8) | -1462 (-2123, -800.5) | -1727 (-2643, -809.9) | -832.0 (-1894, 230.1) | -1321 (-2900, 257.1) | -1251 (-3593, 1092) |

CVD: Cardiovascular Disease; CHD: coronary heart disease; CBVD: cerebrovascular disease; t1: time spline 1; t2: time spline 2; SIMD: Scottish Index of Multiple Deprivation
