## Supplemental tables for "Cost-Effectiveness Analysis of Statins, Berberine, and Their Combined Use for Primary Prevention of Cardiovascular Disease"

Supplemental Table 1. Annual statin and berberine costs from Chinese source

| **Parameters** | **Base case(95%CI)** | **Distribution** | **Data source** | **Reference** |
| --- | --- | --- | --- | --- |
| Intervention cost, £ |  |  |  |  |
| Atorvastatin 20 mg/d | 37.26(8.07~43.81) (¥344.1(74.6~404.6)) | Gamma | Estimate | 48 |
| Berberine 1000mg/d | 26.87(21.96~59.28) (¥248.2(202.8~547.5)) | Gamma | Estimate | 49 |
| Simvastatin 20mg+ berberine 1500mg/d | 49.07(37.11~95.56) (¥450.0(342.6~882.6)) | Gamma | Estimate | 48,49 |
| Simvastatin 20mg+ berberine 900mg/d | 32.23(23.94~59.99) (¥294.6(221.1~554.1)) | Gamma | Estimate | 48,49 |

Supplemental Table 2. Comparison of CVD risk factors for the final intervention-eligible population

| **Variables** | **Overall** | **Male** | **Female** | ***p*** |
| --- | --- | --- | --- | --- |
| n | 3869 | 1718 | 2151 |  |
| Age (years) | 57.13 (11.88) | 56.97 (11.82) | 57.26 (11.94) | 0.460 |
| Survey year, n(%) | |  |  | 0.460 |
| 2003 | 2253 (58.2) | 1006 (58.6) | 1247 (58.0) |  |
| 2008 | 432 (11.2) | 200 (11.6) | 232 (10.8) |  |
| 2009 | 415 (10.7) | 172 (10.0) | 243 (11.3) |  |
| 2010 | 401 (10.4) | 169 (9.8) | 232 (10.8) |  |
| 2011 | 368 (9.5) | 171 (10.0) | 197 (9.2) |  |
| SIMD groups (fifths), n(%) |  |  |  | 0.651 |
| 1 | 764 (19.7) | 337 (19.6) | 427 (19.9) |  |
| 2 | 793 (20.5) | 363 (21.1) | 430 (20.0) |  |
| 3 | 861 (22.3) | 394 (22.9) | 467 (21.7) |  |
| 4 | 784 (20.3) | 339 (19.7) | 445 (20.7) |  |
| 5 | 667 (17.2) | 285 (16.6) | 382 (17.8) |  |
| Diabetes, n(%) |  |  |  | 0.278 |
| No | 3797 (98.1) | 1681 (97.8) | 2116 (98.4) |  |
| Yes | 72 (1.9) | 37 (2.2) | 35 (1.6) |  |
| Family History of CVD, n(%) | | |  | 0.298 |
| No | 3009 (77.8) | 1350 (78.6) | 1659 (77.1) |  |
| Yes | 860 (22.2) | 368 (21.4) | 492 (22.9) |  |
| No. of cigarette per day(n) | 3.37 (7.77) | 3.63 (8.47) | 3.17 (7.15) | 0.064 |
| SBP(mm Hg) | 132.57 (18.66) | 134.52 (16.56) | 131.01 (20.05) | **<0.001** |
| TC(mmol/L) | 5.92 (1.10) | 5.79 (1.07) | 6.03 (1.12) | **<0.001** |
| HDL-c(mmol/L) | 1.54 (0.40) | 1.38 (0.33) | 1.66 (0.40) | **<0.001** |

SIMD: Scottish Index of Multiple Deprivation; CVD: Cardiovascular Disease; SBP: Systolic Blood Pressure; TC: Total Cholesterol; HDL-c: High-Density Lipoprotein Cholesterol

Supplemental Table 3. The cost-effectiveness of Statin, Berberine and Combined Interventions in supplementary analyses

| **Intervention** | **Sample size** | **Number treated** | **Primary CVD events prevented** | **Undiscounted life- years** | **Discounted QALYs gained** | **Discounted cost, £** | **ICER, £/QALY** |
| --- | --- | --- | --- | --- | --- | --- | --- |
| **Intervention strategies comparable to no intervention** | | | | | | | |
| *Intervention eligibility: ASSIGN≥20%* | | |  |  |  |  |  |
| No intervention | 3869 | 0 | Reference | Reference | Reference | Reference | Reference |
| Statins | 3869 | 1331 | 50.2 (32.2~68.5) | 253.2 (68.5~530.1) | 199.3 (54.2~416.9) | 545399.2 (94225.7~1216069.6) | 2736.3 (1355.4~3465.6) |
| Berberine | 3869 | 1331 | 42.1 (28.4~53.4) | 206.6 (76.4~336.1) | 161.5 (58.9~263.9) | 175618.3 (-135836.3~456226.5) | 1087.4 (-2161.6~1849.7) |
| Combined intervention 1 | 3869 | 1331 | 95.4 (68.1~122.3) | 468.8 (107.8~917.2) | 379.7 (98.0~730.0) | 796798.1 (-117758.8~2066011.1) | 2098.6 (-1207.8~2921.3) |
| Combined intervention 2 | 3869 | 1331 | 100.6 (67.6~134.3) | 499.8 (133.3~997.5) | 404.1 (115.8~795.2) | 643938.8 (-233396.9~1949958.8) | 1593.4 (-2025.5~2586.8) |
| *Intervention eligibility: ASSIGN≥10%* | | |  |  |  |  |  |
| No intervention | 3869 | 0 | Reference | Reference | Reference | Reference | Reference |
| Statins | 3869 | 2323 | 89.5 (58.9~120.9) | 431.1 (125.7~877) | 338.7 (97.2~689.9) | 1506324.5 (631948.5~2768509.0) | 4447.0 (3656.3~7032.0) |
| Berberine | 3869 | 2323 | 75.7 (55.4~94.8) | 355.3 (97.1~693.2) | 276.5 (75.1~541.9) | 688692.4 (1572.1~1627619.8) | 2490.8 (36.5~3134.9) |
| Combined intervention 1 | 3869 | 2323 | 171.8 (124.5~217.3) | 803.9 (202.6~1531.4) | 653.8 (180.8~1226.9) | 2349211.6 (623804.3~4622702.7) | 3593.1 (2628.6~4140.1) |
| Combined intervention 2 | 3869 | 2323 | 180.2 (122.3~237.5) | 852.8 (254.0~1672.5) | 692.7 (217.2~1340.0) | 2019913.4 (337149.7~4431792.1) | 2916.1 (1313.3~3465.6) |
| **Intervention strategies comparable to statin intervention** | | | | | | | |
| *Intervention eligibility: ASSIGN≥20%* | | |  |  |  |  |  |
| Statins | 3869 | 1331 | Reference | Reference | Reference | Reference | Reference |
| Berberine | 3869 | 1331 | -8.1 (-15.9~-2.6) | -46.6 (-212.8~49.7) | -37.8 (-167.4~38.6) | -369780.9 (-824741.8~-146924.5) | 9777.9 (-57845.7~74008.7) |
| Combined intervention 1 | 3869 | 1331 | 45.2 (32.7~56.2) | 215.6 (-2.3~431.8) | 180.4 (8.5~347.8) | 251398.9 (-280595.0~886083.6) | 1393.8 (-8397.3~2718.1) |
| Combined intervention 2 | 3869 | 1331 | 50.4 (34.6~66.5) | 246.6 (58.2~477.4) | 204.8 (57.0~385.5) | 98539.6 (-373475.9~758688.3) | 481.1 (-6576.2~1989.7) |
| *Intervention eligibility: ASSIGN≥10%* | | |  |  |  |  |  |
| Statins | 3869 | 2323 | Reference | Reference | Reference | Reference | Reference |
| Berberine | 3869 | 2323 | -13.8 (-26.6~-0.8) | -75.8 (-231.3~37.2) | -62.2 (-186.6~30.3) | -817632.0 (-1253367.0~-522451.4) | 13138.3 (-101288.1~104864.5) |
| Combined intervention 1 | 3869 | 2323 | 82.3 (60.0~101.3) | 372.8 (9.6~726.5) | 315.1 (28.6~592.0) | 842887.1 (-167619.5~1972892.9) | 2675.1 (-1308.4~3471.8) |
| Combined intervention 2 | 3869 | 2323 | 90.7 (63.2~118.5) | 421.7 (107.7~795.7) | 353.9 (107.4~647.0) | 513588.9 (-364956.5~1690371.0) | 1451.0 (-3532.7~2637.1) |

CVD: Cardiovascular Disease; QALY: quality-adjusted life-year; ICER: incremental cost-effectiveness ratio; ICER= Incremental cost / Incremental QALY
