## Supplementary figures and images for "Cost-Effectiveness Analysis of Statins, Berberine, and Their Combined Use for Primary Prevention of Cardiovascular Disease"

### Supplemental figure 1

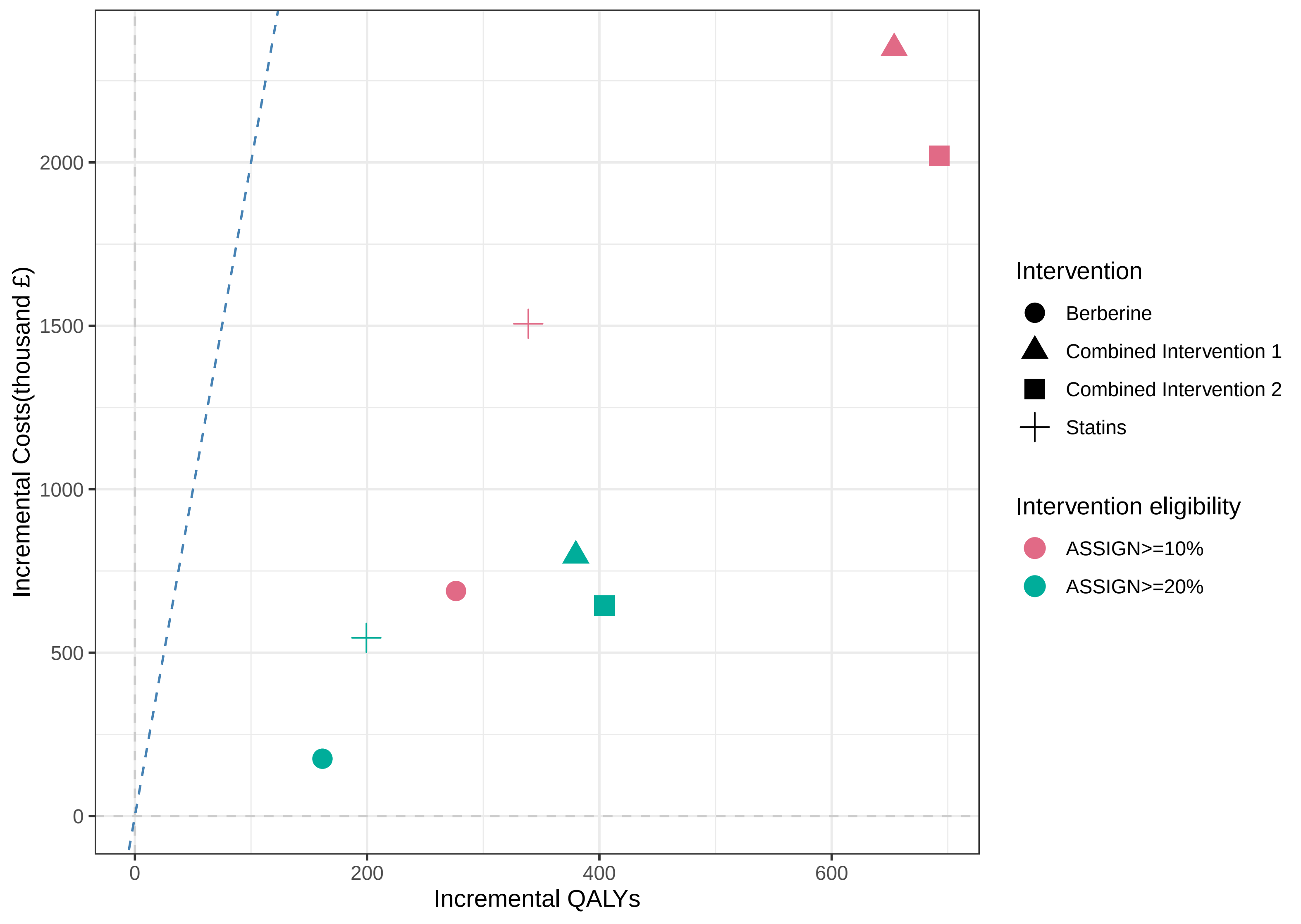

### Supplemental figure 2

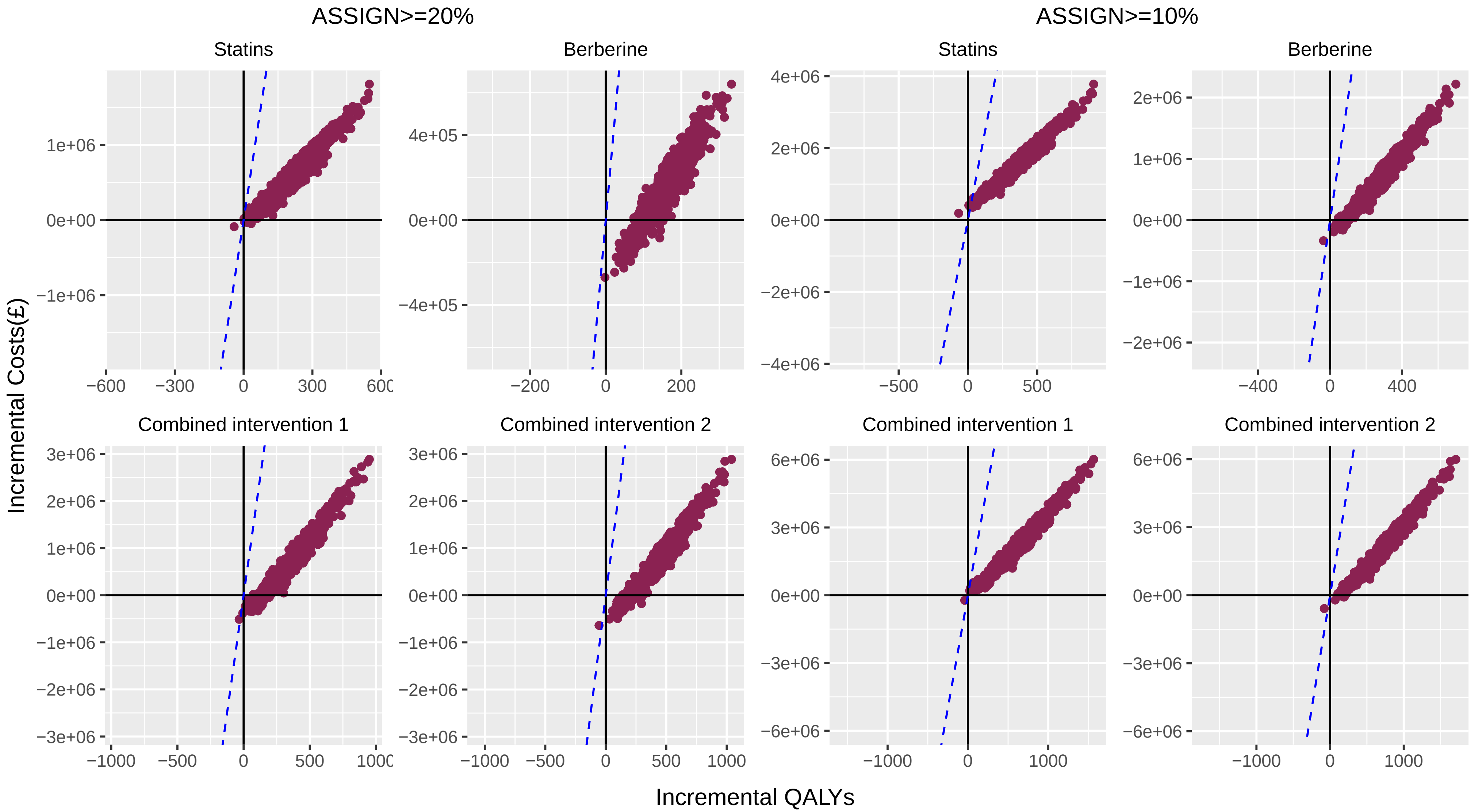

### Supplemental figure 3

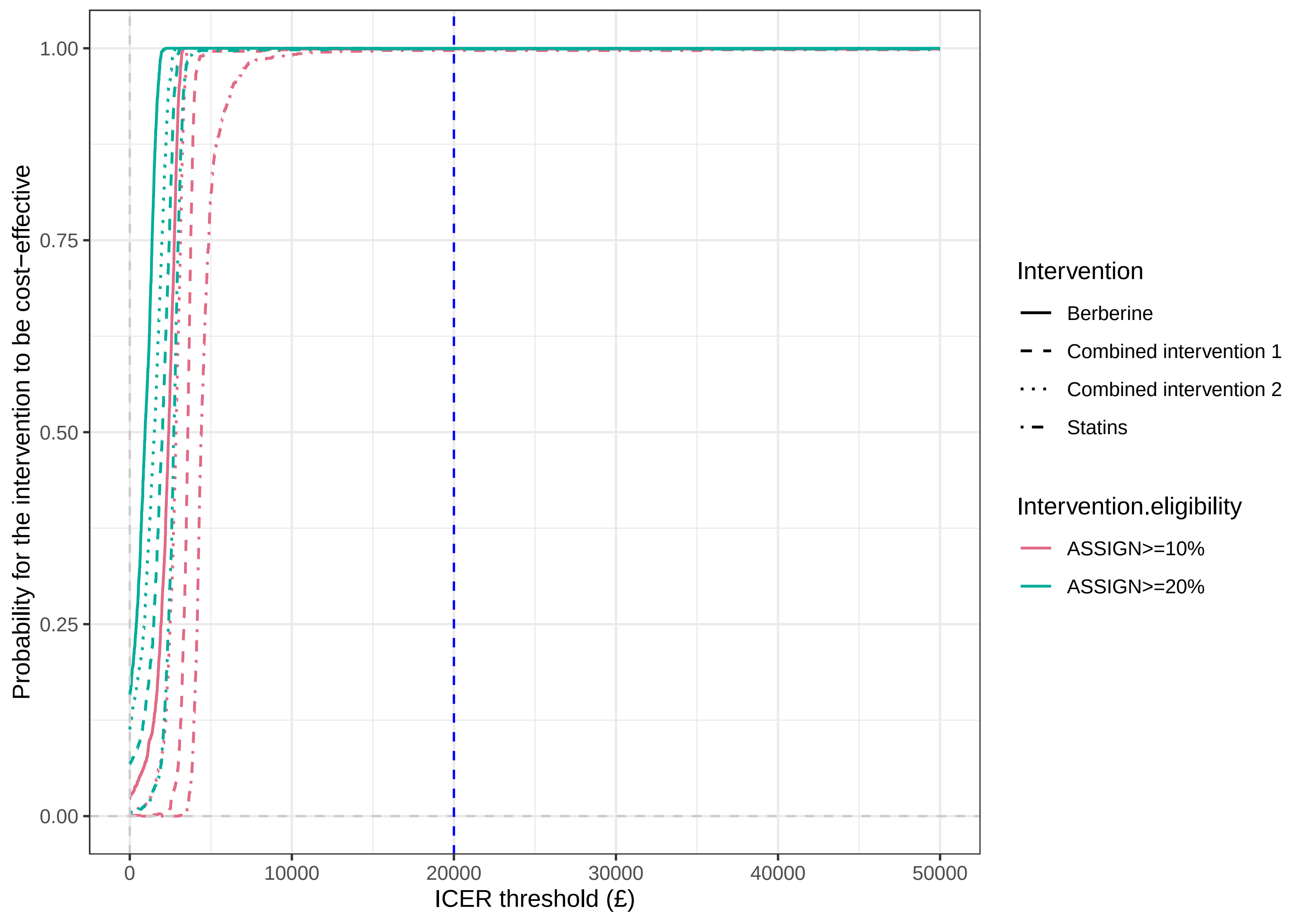
